# Fitting SIR model to COVID-19 pandemic data and comparative forecasting with machine learning

**DOI:** 10.1101/2020.04.26.20081042

**Authors:** Mouhamadou A.M.T. Baldé

## Abstract

In this work, we use a classical SIR model to study COVID-19 pandemic. We aim, to deal with the SIR model fitting to COVID-19 data by using different technics and tools. We particularly use two ways: the first one start by fitting the total number of the confirmed cases and the second use a parametric solver tool. Finally a comparative forecasting, machine learning tools, is given.

## I. Introduction

The Coronavirus disease 2019 (COVID-19) epidemic has to get the attention of all scientists around the world. The COVID-19 appears in November 2019 in Wuhan, central China. In March 2020, the epidemic was reclassified as a pandemic by the World Health Organization (WHO). The COVID-19 pandemic is spreading rapidly in many other countries. Measures, limiting human-to-human contact (social distancing, barrier measures, confinement), are taken to stem the spread, causing a sudden slowdown in the world economy and a stock market crash on March 12, 2020. The COVID-19 is contagious with human-to-human transmission. The incubation period is generally between two and fourteen days, with an average of five days.

Many researchers have used new or classical models of infectious diseases to describe, study or predict the evolution of COVID-19 pandemic.

In this work, we aim to use the well known classical SIR model[Anderson and May, 1991], [Hethcote, 2000], to fit it to world data of COVID-19. We discuss and apply different methods and show numerical results. The methods consist in first to use the work in [Liu, Magal, Seydi and Webb, 2020] which start by fitting the total number of the confirmed cases and in second to use parametric solver.

SIR stands for “Susceptible-Infected-Removed”. If we consider that all removed individuals are the recovered individuals, then we can say “Susceptible-Infected-Recovered”.

In this paper, we first introduce the classical mathematical SIR models we use to fit COVID-19 data. In the second we present different mechanisms we use to fit the data. Then we show numerical results. And finally, we show some prediction for Senegal of the SIR models we study compared with forecasting using machine learning.

## II. classical SIR model

The classical SIR model is given as follow:

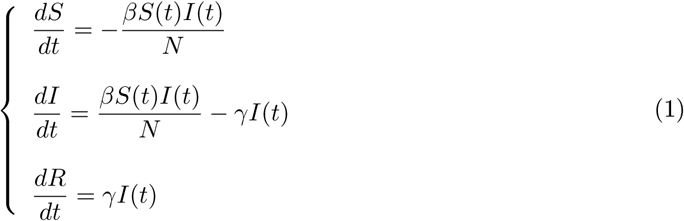

with the dependent variables *S*(*t*), *I*(*t*), *R*(*t*) being respectively the number of susceptible individuals at time *t*, the number of infectious individuals at time *t* and the number of recovered individuals at time *t*. In addition, the unknowns *S*(*t*), *I*(*t*), *R*(*t*) satisfy *N* = *S*(*t*)+*I*(*t*)+*R*(*t*). *N* is the total number of individuals.

The initial conditions are *S*(0) = *S*_0_ ≥ 0; *I*(0) = *I*_0_ ≥ 0 and *R*(0) = *R*_0_ ≥ 0.

Some parameters must be presented: *β* is the contact rate, 1/*γ* is the average infectious period,

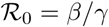 is the basic reproduction number.

The SIR model satisfies some properties:

- The susceptible function *S*(*t*) decreases to 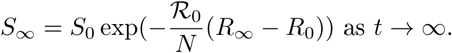
- The recovered function *R*(*t*) increases up to 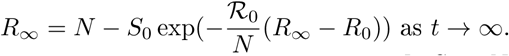
- If 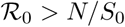, then *I*(*t*) increases up to a maximum value 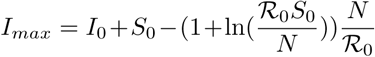.
- If 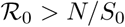, then *I*(*t*) decreases to 0.

### Remark II.1

*At the outset of an epidemic, nearly everyone* (*except the infected case*) *is susceptible. So we can say that S*_0_ ≈ *N and then we can replace N/S_0_ by* 1.

It is also possible to couple the SIR model with other differential equations like death equation to study the number of death due the epidemic disease or economic model equations to study the impact of the epidemic disease on the economy. To couple the SIR model with a death equation, we consider the death rate *μ* due to the infection. So the system is given by:

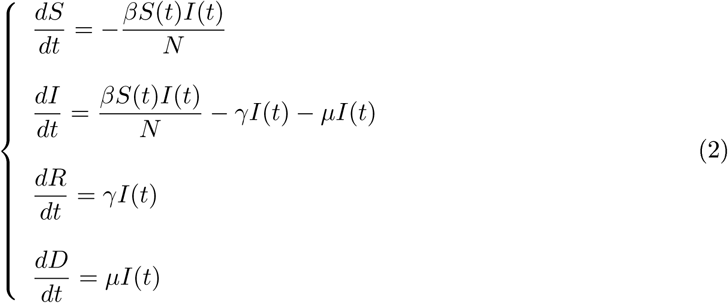

with *D* being the number of death due to the infection. Then the total removed is *R*(*t*) + *D*(*t*) and *N* = *S*(*t*) + *I*(*t*) + *R*(*t*) + *D*(*t*).

## III. Methods

### i. Fitting function to data

The starting point of the work is to find a function that fits the data of total confirmed cases. As in [Liu, Magal, Seydi and Webb, 2020], we set the total number of infectious cases by:

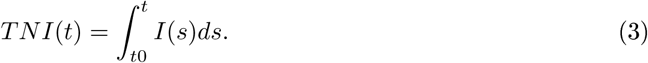

The total number of infectious cases function, that fit the data, can be written as *TNI*(*t*) = *b* exp(*ct*) − *a*. Where *a*, *b*, *c* are parameters to estimate by using the least square method. Then *TNI*(*t*) we can obtain information on the unknown functional variables and parameters of the SIR model.

After calculation as in [Liu, Magal, Seydi and Webb, 2020], we obtain:

- 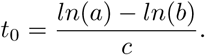
- 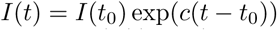, with 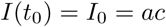.
- 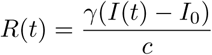.
- 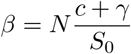.

The simulation results are given in subsection i, figure 1a, 1b, 1c and 1d. Also the values of the parameters *β* and *γ* are given.

**Figure 1:**
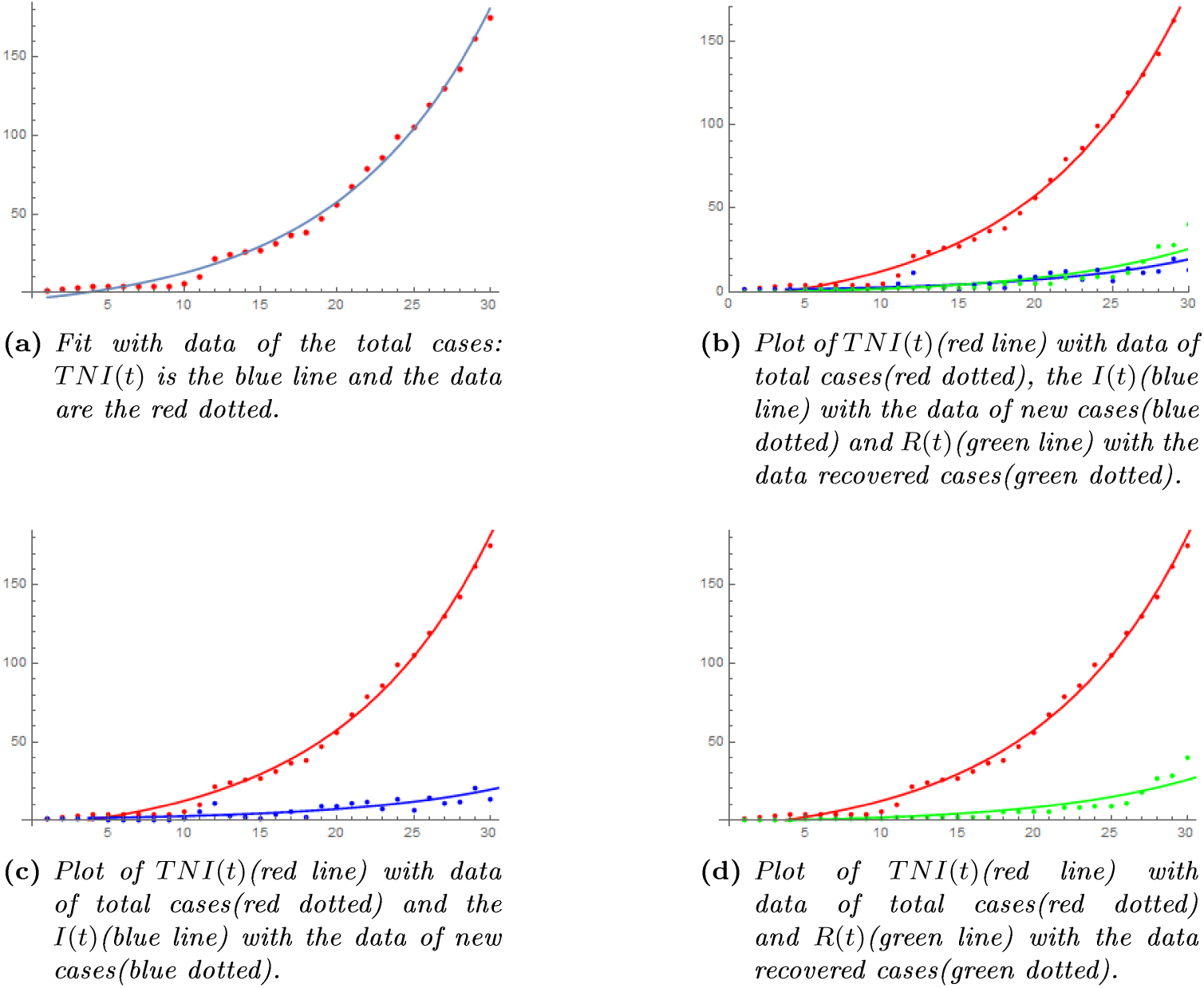
Plot of the total number of case data using the SIR model(1) and compared with plot of Senegal’s data. On the abscissa axis, the graduation 30 represents 2020, March 31.

We consider now that at a time *T* some measures are taken like social distancing, half or full confinement. It can be interpreted as the contact rate is reduced by some factor or at a time point, the contact rate is close to 0. Indeed in [Liu, Magal, Seydi and Webb, 2020], the authors considered that the transmission of COVID-19 from infectious to susceptible individuals stopped after strong measures has been taken in China. Then they have fixed the contact rate to 0. In [Lauro, Kiss and Miller, 2020], the authors considered that the contact rate is reduced by some factor when a social distancing intervention is introduced with a certain duration. Then they have replaced *β* by (1 − *c*)*β*. It is also possible to consider that *β* will decrease depending on time like in [Liu, Magal, Seydi and Webb, 2020], where authors has considered an exponential decreasing function of time.

In this work, we consider that the contact rate decreases progressively to 0. Then we choose continuous function with a slow decrease to describe the contact rate starting at the date-time of the measures.

We propose two types of function for *β* noted 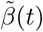.

The first one is:

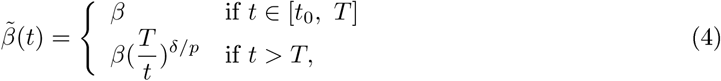

where *δ* and *p* are parameters to choose. The second one is:

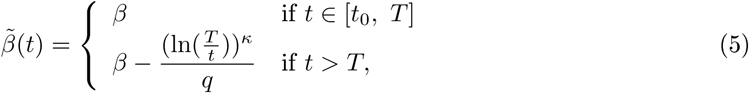

when *κ* and *q* are parameters to choose.

Then replacing *β* by 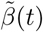 in the SIR model 1, we obtain:

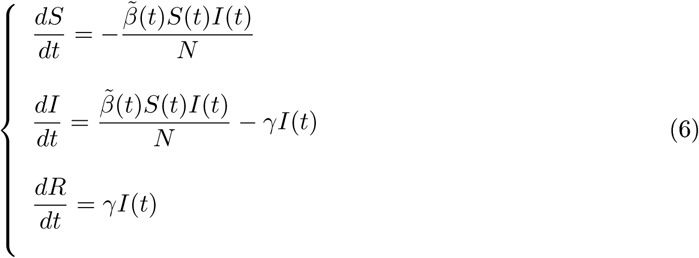

By solving (6), we obtain the new expressions of *TNI*(*t*), *I*(*t*) and *R*(*t*). We show the results in subsection i.

### Remark III.1

1. *The values of the parameters δ, p, κ and, q can be fixed such that* 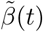 *decreases slowly*.
2. *Since* 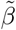 *depends on time, we must always consider values of t such that* 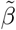 *remains positive*.
3. 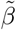 *given by 4 is positive for all time while* 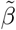 *given by 5 is positive only if t* ∊ [0, *T* exp((*βq*)^1/κ^)].

#### ii. Fit to data with scaling

When there is not enough data the fit is generally difficult since the maximal values of the dependent variables can be very huge. For example, *S*_0_ depends on the total population and generally *S_0_* ≈ *N*, with *N* the size of a chosen sample of the population. In SIR models, it is generally possible to choose different population sizes with similar characteristics. If we have characteristic measures we can make the model dimensionless so that the quality of the results is always good. Then we scale the SIR model to the data by using scaling [Langtangen and Pedersen, 2016].

Let us fix the following constant characteristics size: *t_c_*, *S_c_*, *I_c_*, *R_c_* respectively of the time *t*, the susceptible *S*(*t*), the infected *I*(t) and the recovered *R*(*t*). Then the dimensionless variables 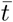, 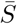, 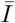, 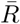 are given by 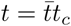, 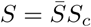, 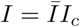, 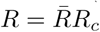.

Replacing in (1) and calculating, we obtain the scaled system:

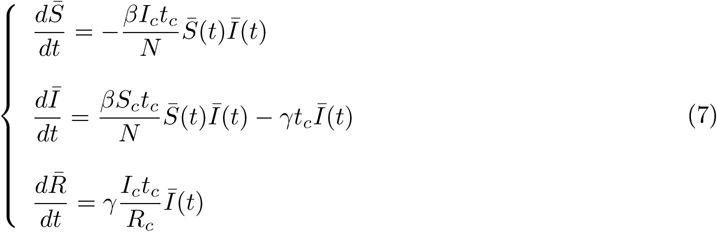

### Remark III.2

For the adjusting of the data, we choose 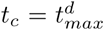, with 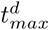 being the last of the data.

The scaled system can be written, by dropping “-” as follows:

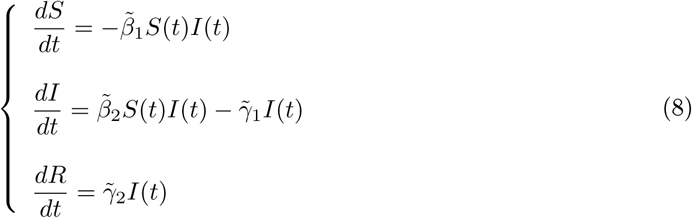

with 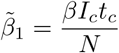, 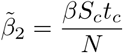, 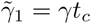 and 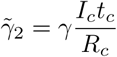.

### Remark III.3

*By choosing the characteristics such that S_c_ = I_c_ = R_c_, we obtain* 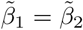 *and*, 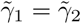. *And then vie get a new SIR model*.

#### iii. Parametric solve

We estimate the parameters *β* mid *γ* by using parametric solver in Mathematica. With the parametric solver, we solve the SIR model (2) with solutions depending on the parameters we want to estimate such that the model fits the data. Then plotting the parametric solution for different values of *β* and *γ*, finally, give a fit. Parametric solver typically solve differential equations by going through several different stages, depending on the type of equations.

#### iv. Machine learning

Machine learning is programming computers to optimize a performance criterion using example data or past experience. We have a model defined up to some parameters, and learning is the execution of a computer program to optimize the parameters of the model using the training data or past experience. The model may be predictive to make predictions in the future, or descriptive to gain knowledge from data or both.

Machine learning uses the theory of statistics in building mathematical models because the core task is making inferences from a sample. (See [Alpaydin, 2010]).

Then machine learning can be used for the estimation of parameters of an epidemic model. We can go deep by doing data mining since we have massive world data. We can use machine learning to learn about the worldwide data of pandemic COVID-19 and then to predict the future evolution of the disease.

We can also try to learn on the spatial distribution and progression of the disease and predict the location susceptible to be a high level of risk.

In this work, we just use a machine learning tool to learn on small data of Senegal’s COVID-19 cases and then to predict the evolution in future days. We use it as a comparison with the other forecasts based on the work we do in this paper.

### IV. Numerical simulations

#### i. The data

In this subsection we show table of data we use for simulation. The tables 1, 2 and 3 give data respectively of Senegal, China and France. Data for Senegal is obtained from daily press releases on the COVID-19 from the Ministry of Health and Social Action (http://www.sante.gouv.sn/) and the data for China and France come from [Wolfram Research]. [Wolfram Research] provide estimated confirmed COVID-19 infection trend by country or region, based on WHO, U.S. CDC, ECDC, China CDC (CCDC), NHC and DXY. The source dataset is compiled daily by Johns Hopkins CSSE. For details see: https://github.com/CSSEGISandData/COVID-19/issues.

**Table 1:** COVID-19 data for Senegal: 2020 march 02-31.

| Date | 2 | 3 | 4 | 5 | 6 | 7 | 8 | 9 | 10 | 11 | 12 | 13 | 14 | 15 | 16 | 17 | 18 | 19 | 20 | 21 | 22 | 23 | 24 | 25 | 26 | 27 | 28 | 29 | 30 | 31 |
| --- | --- | --- | --- | --- | --- | --- | --- | --- | --- | --- | --- | --- | --- | --- | --- | --- | --- | --- | --- | --- | --- | --- | --- | --- | --- | --- | --- | --- | --- | --- |
| New cases | 1 | 1 | 1 | 1 | 0 | 0 | 0 | 0 | 0 | 1 | 5 | 11 | 3 | 2 | 1 | 4 | 5 | 2 | 9 | 9 | 11 | 12 | 7 | 13 | 6 | 14 | 11 | 12 | 20 | 13 |
| Total cases | 1 | 2 | 3 | 4 | 4 | 4 | 4 | 4 | 4 | 5 | 10 | 21 | 24 | 26 | 27 | 31 | 36 | 38 | 47 | 56 | 67 | 79 | 86 | 99 | 105 | 119 | 130 | 142 | 162 | 175 |
| Recovered cases | 0 | 0 | 0 | 0 | 1 | 1 | 1 | 1 | 1 | 1 | 2 | 2 | 2 | 2 | 2 | 2 | 2 | 5 | 5 | 5 | 5 | 8 | 8 | 9 | 9 | 11 | 18 | 27 | 28 | 40 |
| Death cases | 0 | 0 | 0 | 0 | 0 | 0 | 0 | 0 | 0 | 0 | 0 | 0 | 0 | 0 | 0 | 0 | 0 | 0 | 0 | 0 | 0 | 0 | 0 | 0 | 0 | 0 | 0 | 0 | 0 | 1 |

**Table 2:** COVID-19 data for Hubei, China: 2020 January 22-Marsh 31.

| Date | 1 | 2 | 3 | 4 | 5 | 6 | 7 | 8 | 9 | 10 | 11 | 12 | 13 | 14 | 15 | 16 | 17 | 18 |
| --- | --- | --- | --- | --- | --- | --- | --- | --- | --- | --- | --- | --- | --- | --- | --- | --- | --- | --- |
| Infected cases | 399 | 399 | 494 | 689 | 964 | 1302 | 3349 | 3341 | 4651 | 5461 | 6736 | 10532 | 12722 | 15677 | 18483 | 20677 | 23139 | 24881 |
| Recovered cases | 28 | 28 | 31 | 32 | 42 | 45 | 80 | 88 | 90 | 141 | 168 | 295 | 386 | 522 | 633 | 817 | 1115 | 1439 |
| Death cases | 17 | 17 | 24 | 40 | 52 | 76 | 125 | 125 | 162 | 204 | 249 | 350 | 414 | 479 | 549 | 618 | 699 | 780 |
| Date | 19 | 20 | 21 | 22 | 23 | 24 | 25 | 26 | 27 | 28 | 29 | 30 | 31 | 32 | 33 | 34 | 35 |  |
| Infected cases | 26965 | 28532 | 29659 | 29612 | 43437 | 48175 | 49030 | 49847 | 50338 | 50633 | 49665 | 48510 | 48637 | 46439 | 46395 | 45044 | 43252 |  |
| Recovered cases | 1795 | 2222 | 2639 | 2686 | 3459 | 4774 | 5623 | 6639 | 7862 | 9128 | 10337 | 11788 | 11881 | 15299 | 15343 | 16748 | 18971 |  |
| Death cases | 871 | 974 | 1068 | 1068 | 1310 | 1457 | 1596 | 1696 | 1789 | 1921 | 2029 | 2144 | 2144 | 2346 | 2346 | 2495 | 2563 |  |
| Date | 36 | 37 | 38 | 39 | 40 | 41 | 42 | 43 | 44 | 45 | 46 | 47 | 48 | 49 | 50 | 51 | 52 |  |
| Infected cases | 41603 | 39572 | 36829 | 34617 | 32610 | 30366 | 28174 | 25904 | 23972 | 22628 | 21207 | 19486 | 18247 | 16993 | 15593 | 14407 | 13171 |  |
| Recovered cases | 20969 | 23383 | 26403 | 28993 | 31536 | 33934 | 36208 | 38557 | 40592 | 42033 | 43500 | 45235 | 46488 | 47743 | 49134 | 50318 | 51553 |  |
| Death cases | 2615 | 2641 | 2682 | 2727 | 2761 | 2803 | 2835 | 2871 | 2902 | 2931 | 2959 | 2986 | 3008 | 3024 | 3046 | 3056 | 3062 |  |
| Date | 53 | 54 | 55 | 56 | 57 | 58 | 59 | 60 | 61 | 62 | 63 | 64 | 65 | 66 | 67 | 68 | 69 | 70 |
| Infected cases | 11755 | 10421 | 9557 | 8685 | 7751 | 6988 | 6285 | 5715 | 5223 | 4765 | 4317 | 3827 | 3431 | 2895 | 2526 | 2049 | 1726 | 1461 |
| Recovered cases | 52960 | 54288 | 55142 | 56003 | 56927 | 57682 | 58382 | 58946 | 59433 | 59882 | 60324 | 60811 | 61201 | 61732 | 62098 | 62570 | 62889 | 63153 |
| Death cases | 3075 | 3085 | 3099 | 3111 | 3122 | 3130 | 3133 | 3139 | 3144 | 3153 | 3160 | 3163 | 3169 | 3174 | 3177 | 3182 | 3186 | 3187 |

#### ii. The function fit

Here we show simulations related to the subsection i of section III. The formula of *TNI*(*t*) is given by *TNI*(*t*) = *b* exp(*ct*) − *a*, with *a* = 13.9324, *b* = 9.61779 and *c* = 0.100095 (figure la). We use *γ* = 1/7 and then we obtain: *t*_0_ = 3.7051, *I*_0_ = 1,39456. The total population of Senegal is *N* = 16743927 from Senegal Population (2020) - Worldometer (www.worldometers.info) and then we obtain *S_0_* = *N* − *I*_0_. Sinee *S_0_* ≈ *N* we simplify the calculation and obtain *β* = 0, 242957.

Now we consider the time of the measures as 2020, March 23. Then *T* = 23. For 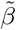 given by (4), results are shown by figures 2a, 2c and 2e. For 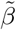 given by (5), results are shown by figures 2b, 2d and 2f.

**Figure 2:**
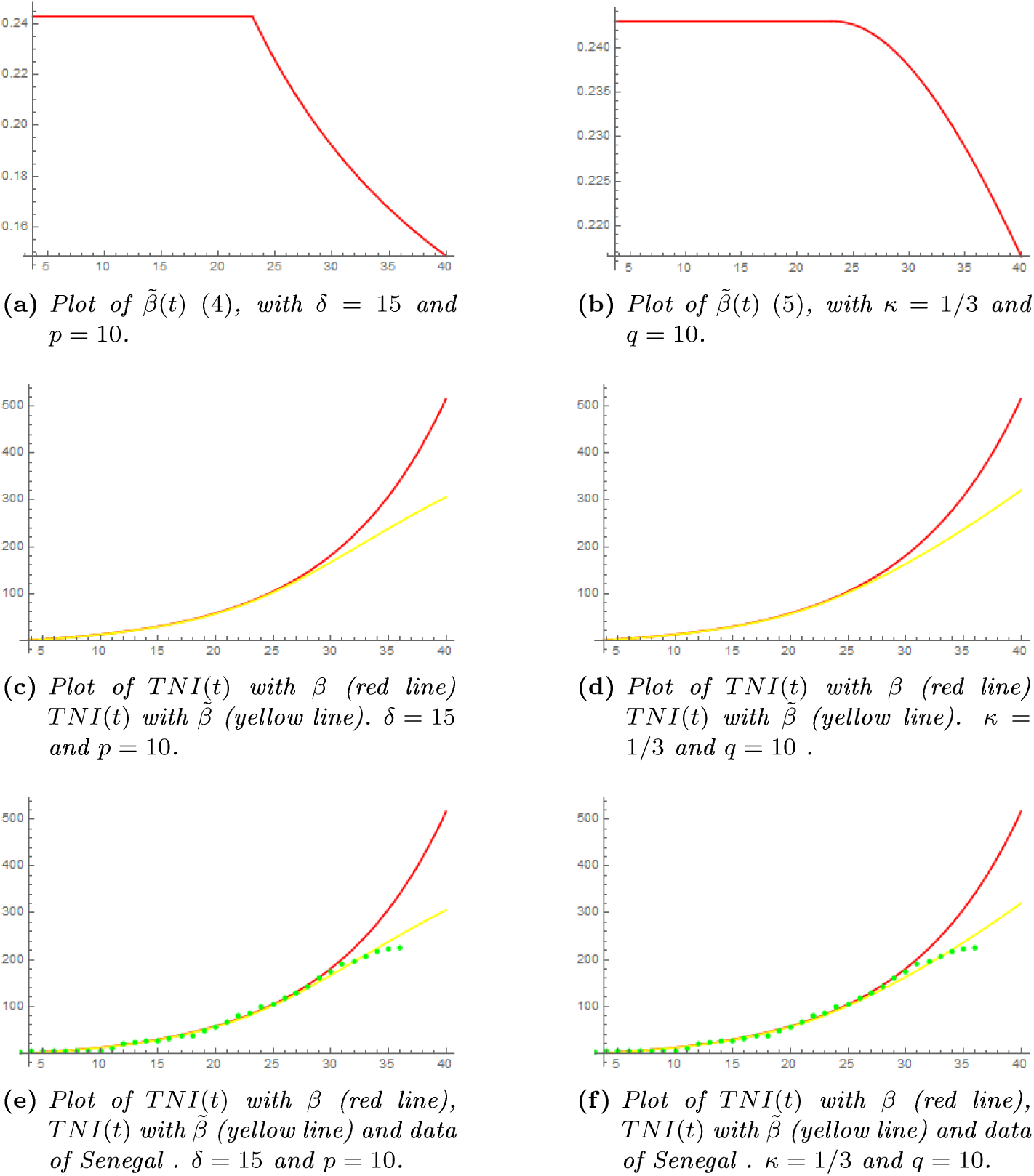
On the top: plot of the contact rate 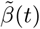 (4) with, δ = 15 and p = 10 and 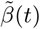 (5) with, κ = 1/3 and q = 10. In the left for figures related to 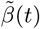 (4) and in the right for figures related to 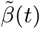 (5); we plot the two functions TNI(t) compared with, plot of Senegal’s data in table 1 completed until April 06. On the abscissa axis, the graduation 30 represents 2020, March 31. Then 2020, April 06 is the graduation 36.

**Table 3:** COVID-19 data for France: 2020 January 22-Marsh 31.

|  |  |  |  |  |  |  |  |  |  |  |  |  |  |  |  |  |  |  |
| --- | --- | --- | --- | --- | --- | --- | --- | --- | --- | --- | --- | --- | --- | --- | --- | --- | --- | --- |
| Date | 1 | 2 | 3 | 4 | 5 | 6 | 7 | 8 | 9 | 10 | 11 | 12 | 13 | 14 | 15 | 16 | 17 | 18 |
| Infected cases | 0 | 0 | 2 | 3 | 3 | 3 | 4 | 5 | 5 | 5 | 6 | 6 | 6 | 6 | 6 | 6 | 6 | 11 |
| Recovered cases | 0 | 0 | 0 | 0 | 0 | 0 | 0 | 0 | 0 | 0 | 0 | 0 | 0 | 0 | 0 | 0 | 0 | 0 |
| Death cases | 0 | 0 | 0 | 0 | 0 | 0 | 0 | 0 | 0 | 0 | 0 | 0 | 0 | 0 | 0 | 0 | 0 | 0 |
| Date | 19 | 20 | 21 | 22 | 23 | 24 | 25 | 26 | 27 | 28 | 29 | 30 | 31 | 32 | 33 | 34 | 35 |  |
| Infected cases | 11 | 11 | 11 | 9 | 9 | 9 | 7 | 7 | 7 | 7 | 7 | 7 | 7 | 7 | 7 | 7 | 2 |  |
| Recovered cases | 0 | 0 | 0 | 2 | 2 | 2 | 4 | 4 | 4 | 4 | 4 | 4 | 4 | 4 | 4 | 4 | 11 |  |
| Death cases | 0 | 0 | 0 | 0 | 0 | 0 | 1 | 1 | 1 | 1 | 1 | 1 | 1 | 1 | 1 | 1 | 1 |  |
| Date | 36 | 37 | 38 | 39 | 40 | 41 | 42 | 43 | 44 | 45 | 46 | 47 | 48 | 49 | 50 | 51 | 52 |  |
| Infected cases | 5 | 25 | 44 | 86 | 116 | 176 | 188 | 269 | 359 | 632 | 926 | 1095 | 1178 | 1739 | 2221 | 2221 | 3570 |  |
| Recovered cases | 11 | 11 | 11 | 12 | 12 | 12 | 12 | 12 | 12 | 12 | 12 | 12 | 12 | 12 | 12 | 12 | 12 |  |
| Death cases | 2 | 2 | 2 | 2 | 2 | 3 | 4 | 4 | 6 | 9 | 11 | 19 | 19 | 33 | 48 | 48 | 79 |  |
| Date | 53 | 54 | 55 | 56 | 57 | 58 | 59 | 60 | 61 | 62 | 63 | 64 | 65 | 66 | 67 | 68 | 69 | 70 |
| Infected cases | 4366 | 4396 | 6473 | 7492 | 8883 | 10616 | 12150 | 13708 | 13144 | 16796 | 17923 | 20002 | 22511 | 25269 | 29561 | 30366 | 33599 | 39161 |
| Recovered cases | 12 | 12 | 12 | 12 | 12 | 12 | 12 | 12 | 2200 | 2200 | 3281 | 3900 | 4948 | 5700 | 5700 | 7202 | 7927 | 9444 |
| Death cases | 91 | 91 | 148 | 148 | 148 | 243 | 450 | 562 | 674 | 860 | 1100 | 1331 | 1696 | 1995 | 2314 | 2606 | 3024 | 3523 |

We do the same as in the paragraphs above for France. *TNI*(*t*) = *b* exp(*ct*) − *a*, with *a* =1, *b* = 0.88 and *c* = 0.158 (figure 3a). We use *γ* = 1/7 and then we obtain: *t*_0_ = 0.809, *I*_0_ = 0.158. The total population of France is *N* = 65241903 from France Population 2020 (https://worldpopulationreview.com) and then we obtain *S*_0_ = *N* − *I*_0_. Since *S*_0_ ≈ *N* we simplify the calculation and obtain *β* = 0.300857.

**Figure 3:**
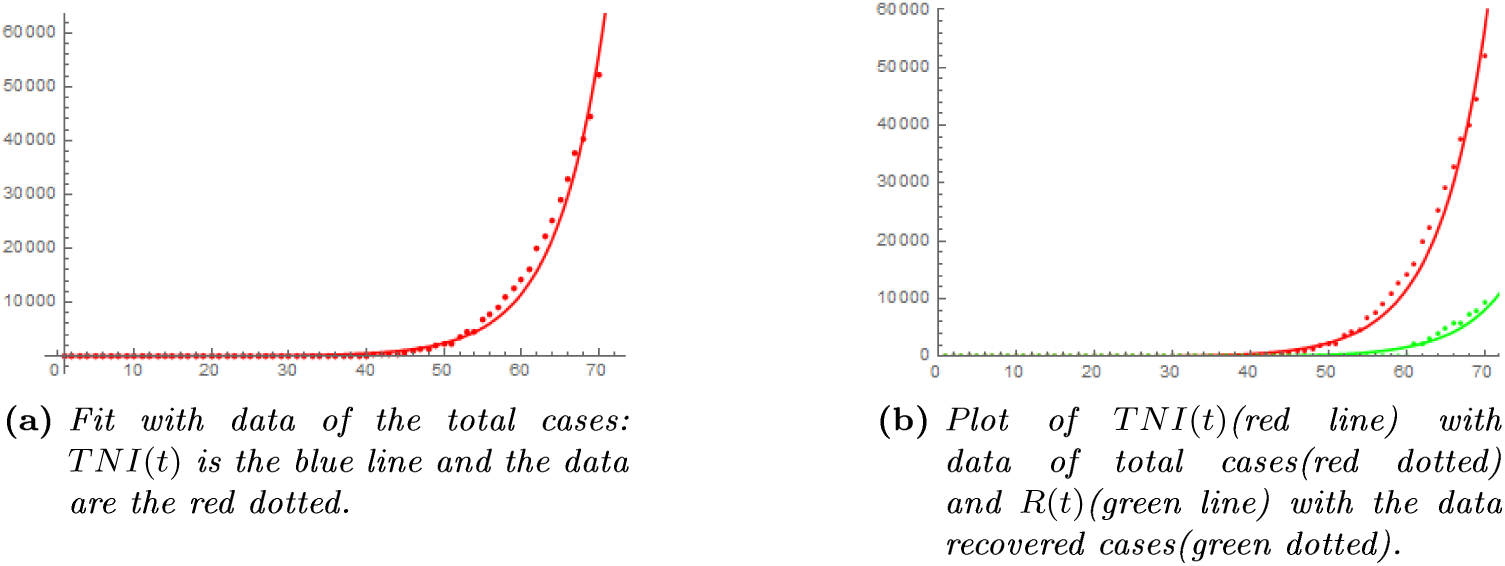
Plot of the total number of case data using the SIR model (1) and compared with plot of France’s data. On the abscissa axis, the graduation 70 represents 2020, March 31.

For France, we consider the time of the measures as 2020, March 17. Then *T* = 56. For 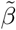 given by (4), results are shown by figures 4a and 4b. For 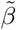 given by (5), results are shown by figures 5a and 5b.

**Figure 4:**
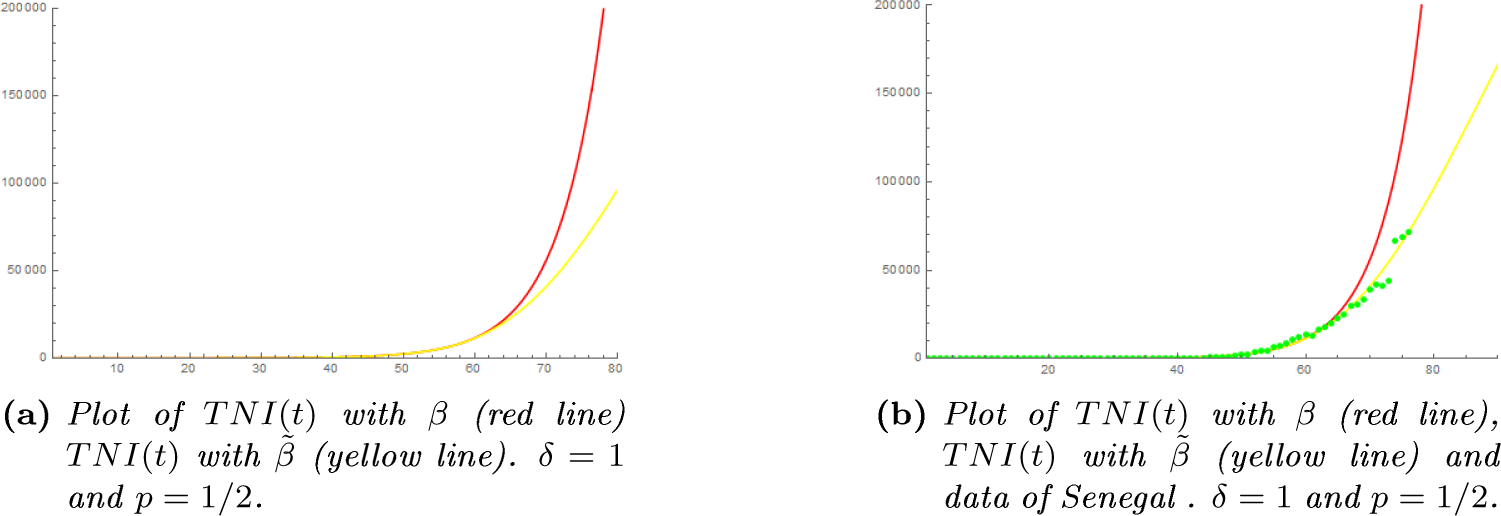
Plot of the contact rate 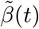 (4) with, δ = 1 and p = 1/2. Plot of the two functions TNI(t) compared with plot of France’s data in table 3 completed until April 06. On the abscissa axis, the graduation 70 represents 2020, March 31.

**Figure 5:**
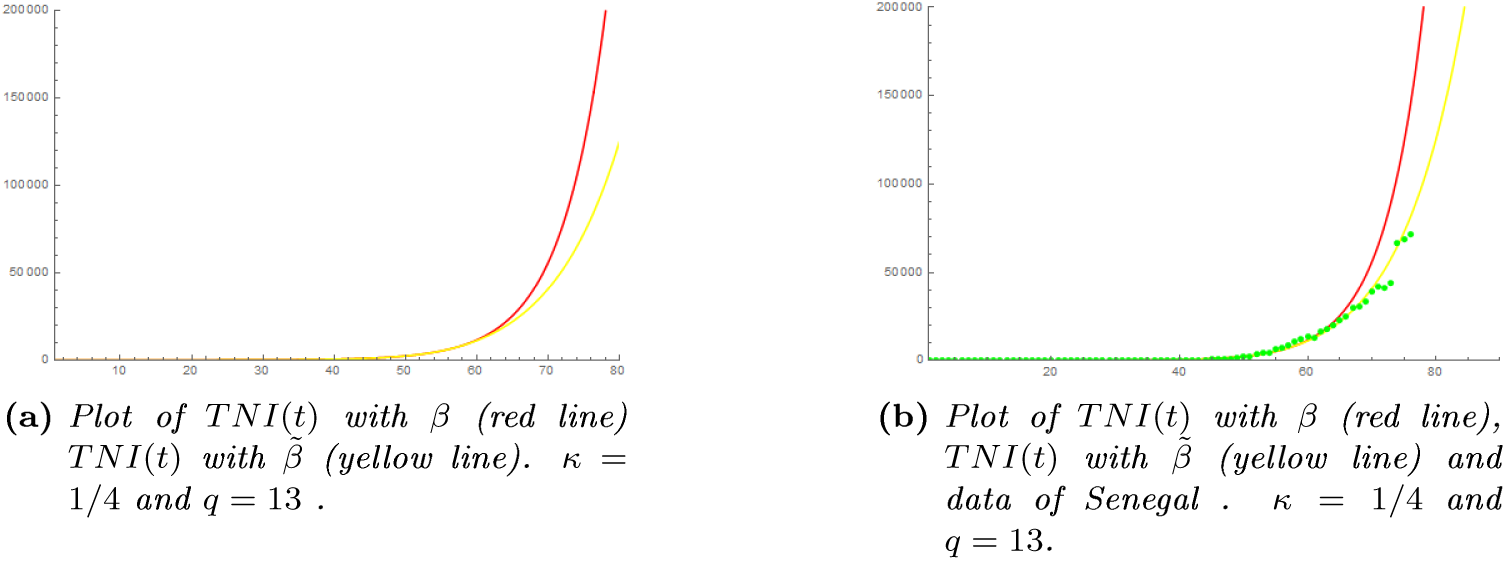
Plot of the contact rate 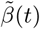 (5) with, κ = 1/4 and q = 13. Plot the two functions TNI(t) compared with plot of France’s data in table 1 completed until April 06. On the abscissa axis, the graduation 70 represents 2020, March 31. Then 2020, April 06 is the graduation 76.

#### iii. The fit by scaling

The scaled SIR model (8) is plotted in figure 6a. And in figure 6b we fit with the data of Senegal in table 1. We fix the characteristic parameters as: *t_c_* = 30 which is the number of days of the data of Senegal in table 1, *I_c_* = *S_c_* = *R_c_* = 1000. We use *β* = 5.0753, *γ* = 1/45 to obtain *β*_1_ = *β*_2_ = 0.00939349 and *γ*_1_ = *γ*_2_ = 0.535714.

**Figure 6:**
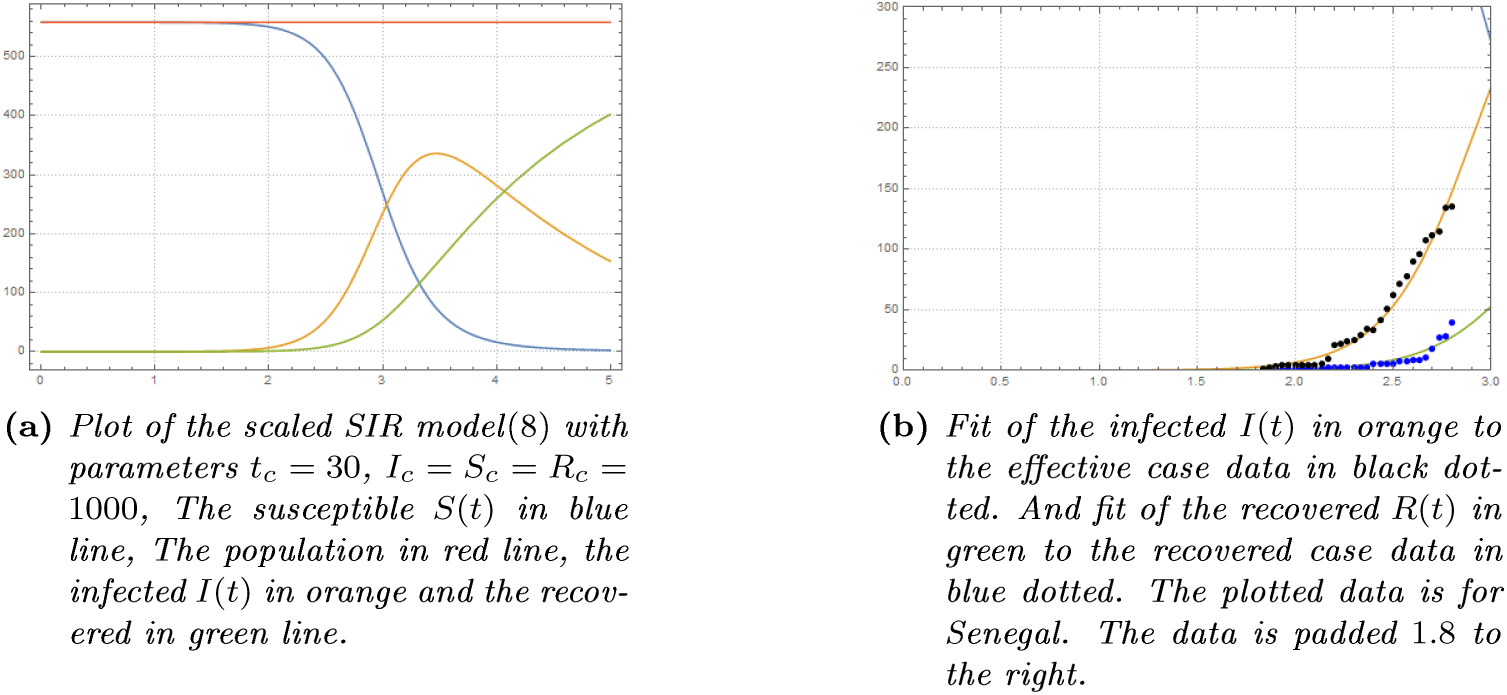
Plot of the scaled SIR model(8) compared with plot of Senegal’s data in table 1. The date 2020 March 31 corresponds to the graduation 2.8.

#### iv. The fit by parametric resolution

Here we use a parametric solver in Wolfram Mathematica to solve the SIR model (2) with respect to parameters *β* and *γ*. We start % using the values *β* = 5.0753 and *γ* = 1/45. Then after we get the new values for the fit. The results are shown in figures 7a. 7b and 7c for Senegal, in figures 8a. 8b and 8c for China and in figures 9a. 9b and 9c for France.

**Figure 7:**
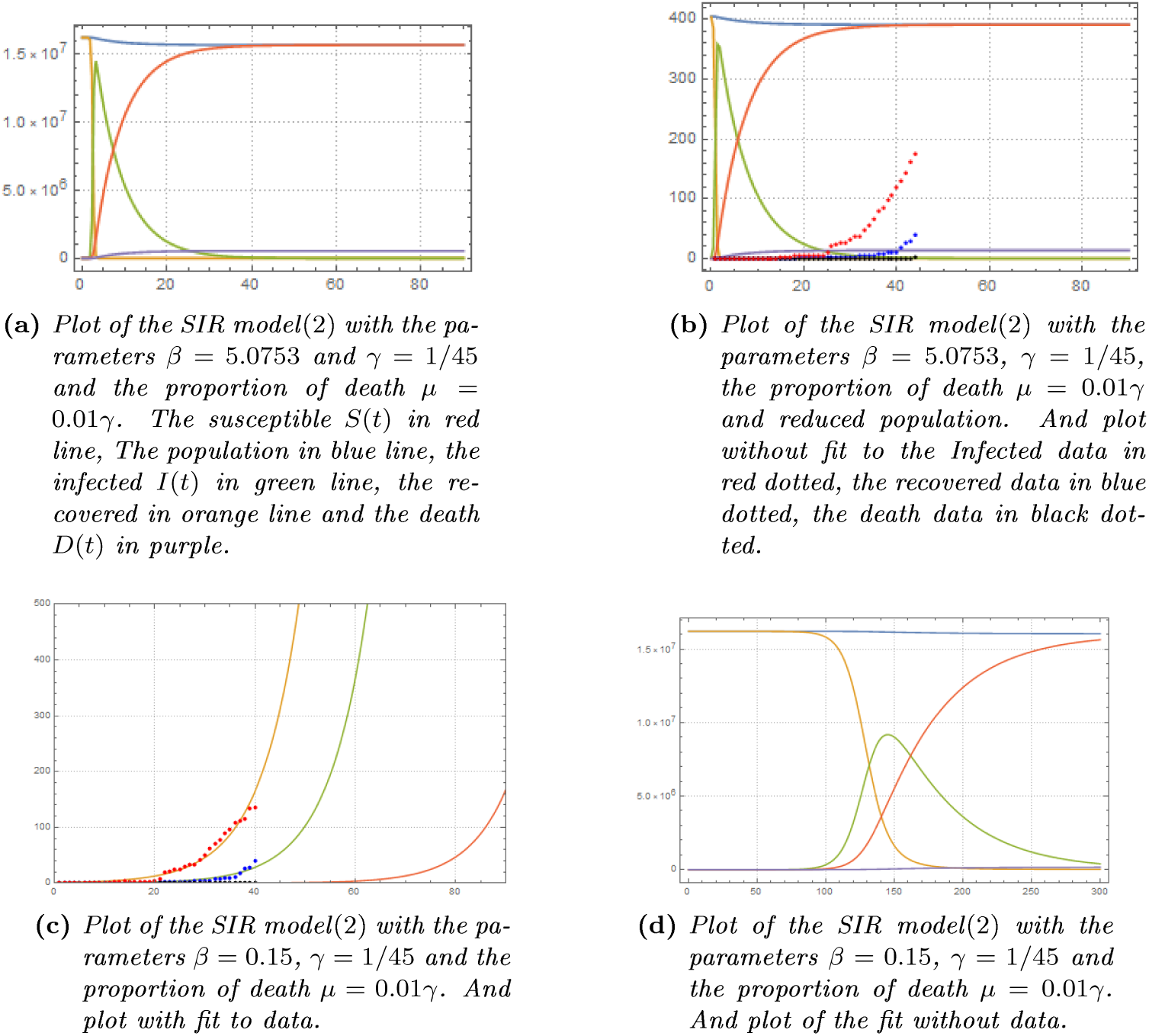
Plot of the SIR model(2) compared with plot of Senegal’s data in table 1. The total population of Senegal we use is N=16743927. On the abscissa axis, the graduation 40 represents 2020, March 31.

**Figure 8:**
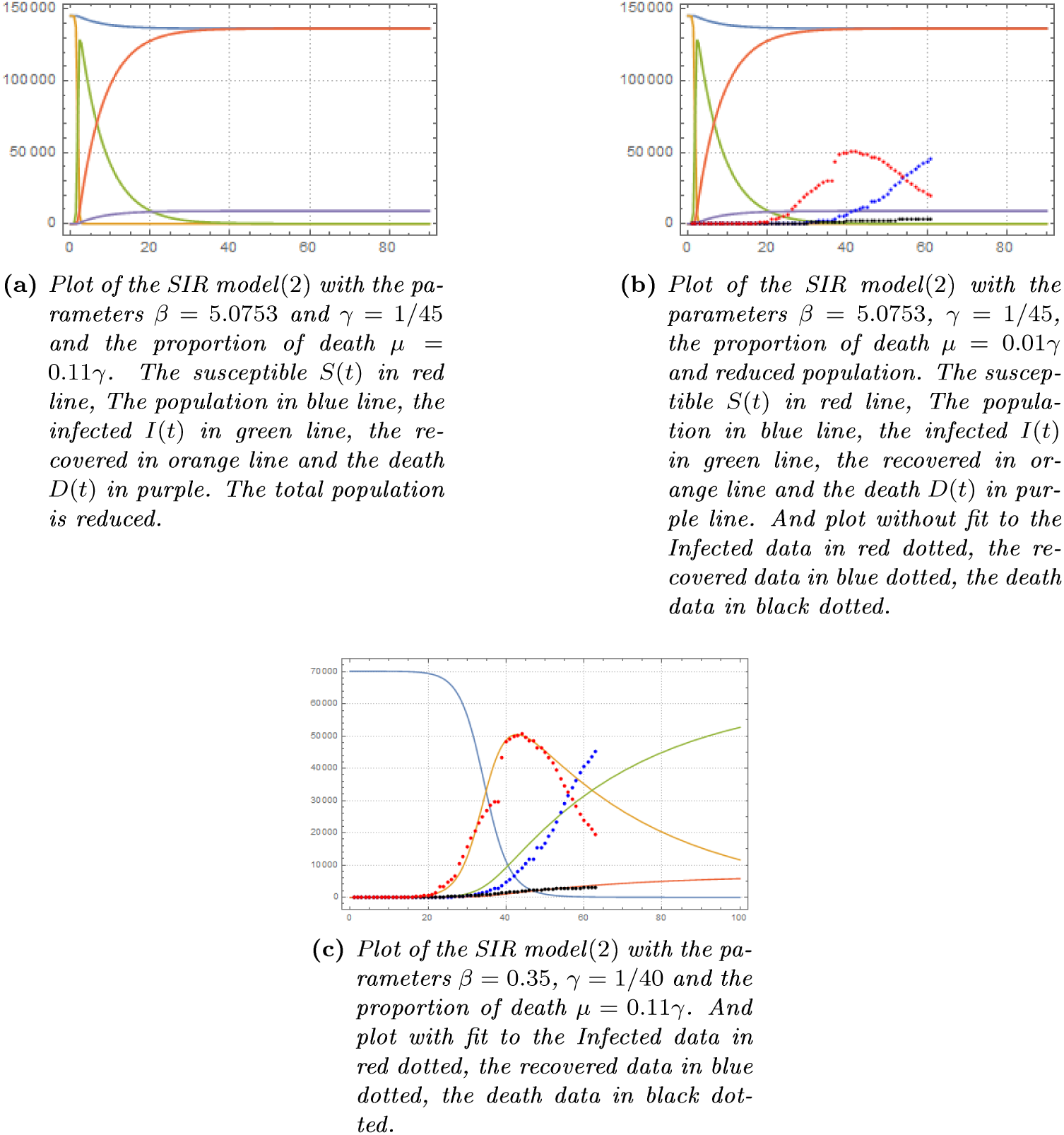
Plot of the SIR model(2) compared with plot of China’s data in table 2. The total population of Hubei we use is N=59172000 (https://en.mkipedia.org/wiki/2020_Hubei_lockdowns). On the abscissa axis, the graduation 63 represents 2020, March 08.

**Figure 9:**
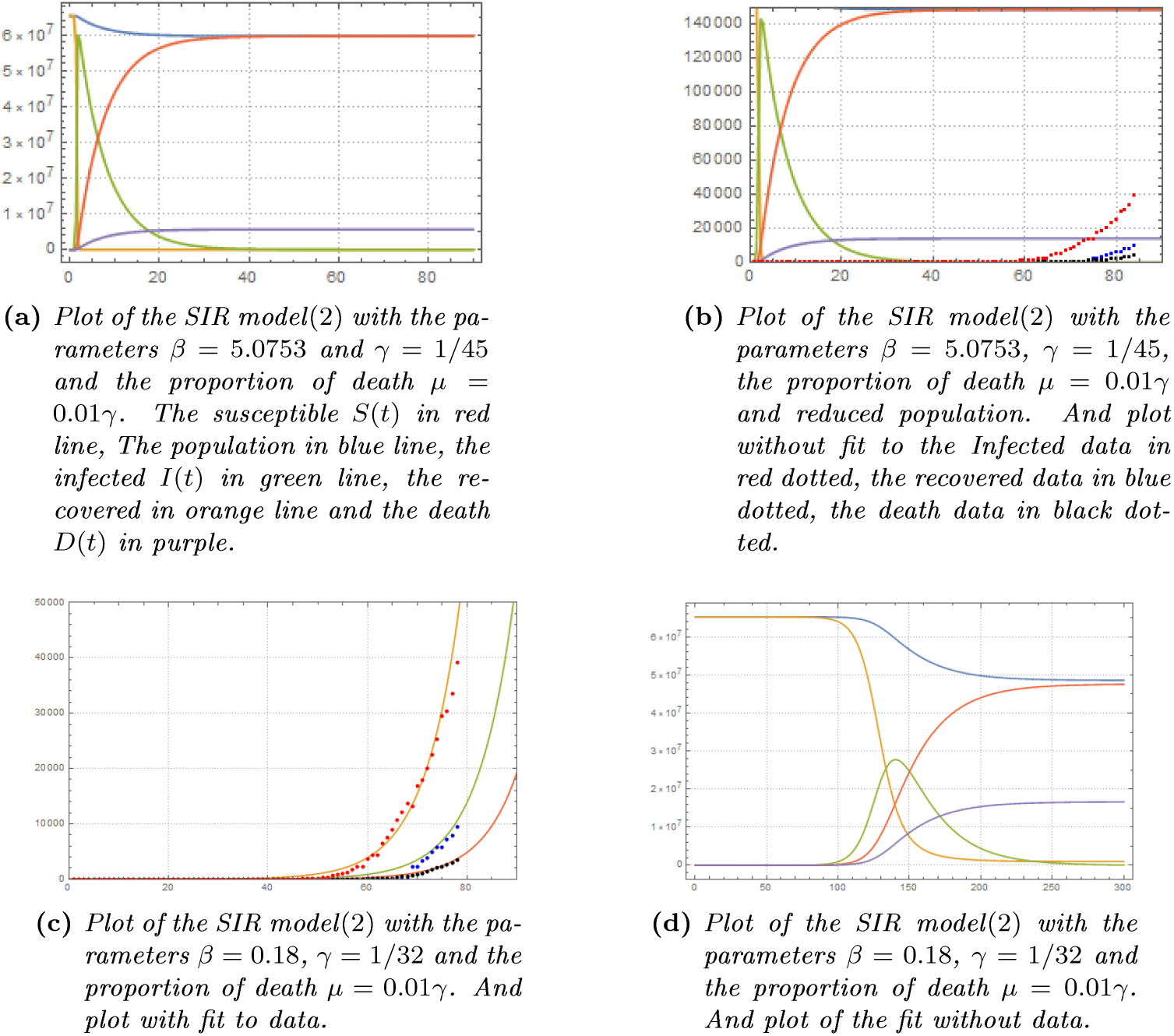
Plot of the SIR model(2) compared with plot of France’s data in table 3. The total population of France we use is N=65241903. On the abscissa axis, the graduation 63 represents 2020, March 31.

### v. Discussion

The SIR model is fitted to the data. This allows us to note that the maximum number of infected can go up to 900000 (figure 7d) in Senegal and 2700000 (figure 9d) in France. While in the scaling the figures 6a shows that the maximum number of infected can go up to 320000. However, this analysis does not take into account the nationwide anti-pandemic measures.

Using the results shown in the figures 2c, 2e, 2d, 2f, 4a, 4b, 5a and 5b, we can see that the data deviate from their first exponential evolution to follow a new, slower trajectory which can still be exponential. To understand the nature of this new trajectory, the way to manage the contact rate is decisive and can lead to different analyzes. However, having considered a slow decrease over time of the contact rate, we can see in the figures 2e, 2f, 4b and 5b that the updated data pass under the new trajectory for Senegal while in France the data follow the new trajectory. This could, therefore, be due to the effects of the measures taken by these countries.

## V. Prediction

In this section, we plot some predictions for Senegal. The prediction does not concern the total confirmed cases, but only the effective cases obtained by reduced from the total confirmed cases, the recovered cases and the death cases.

We show the curve of effective infected cases given by *TNI*(*t*) − *R*(*t*) − *D*(*t*) with both 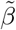 (4) and (5), the curve of the fitted infected *I*(*t*) in figure 7c, and the curve obtained by using machine learning. We plot these curves with additional dates until 2020, April 21.

We use machine learning, based only on data, to do forecasting.

“Predict” is a function of “Automated Machine Learning” in Wolfram Mathematica. It allows for automatic training and data prediction. We can choose different method of regression algorithm: “RandomForest”, “LinearRegression”, “NeuralNetwork”, “GaussianProcess”, “NearestNeighbors”, etc. We use the “NeuralNetwork” regression algorithm which predicts using an artificial neural network.

Let’s recall that our forecasting use in a first part the results of the works in subsection i,ii and iii where we use a SIR model to fit data. And in a second part, the forecasting using only data. The forecasting show in figures 10a, 10c an optimistic situation for Senegal. Also the machine learning figure 10e shows an optimistic forecasting. But in all cases, we see that the additional data (March 31-06) goes down the predicting path. It may be caused by the nationwide anti-pandemic measures in Senegal. In addition, we have considered in figures 10a and 10b a contact rate gradually reduced since the measures were taken and we see that the data come below the path of the predicted curve.

**Figure 10:**
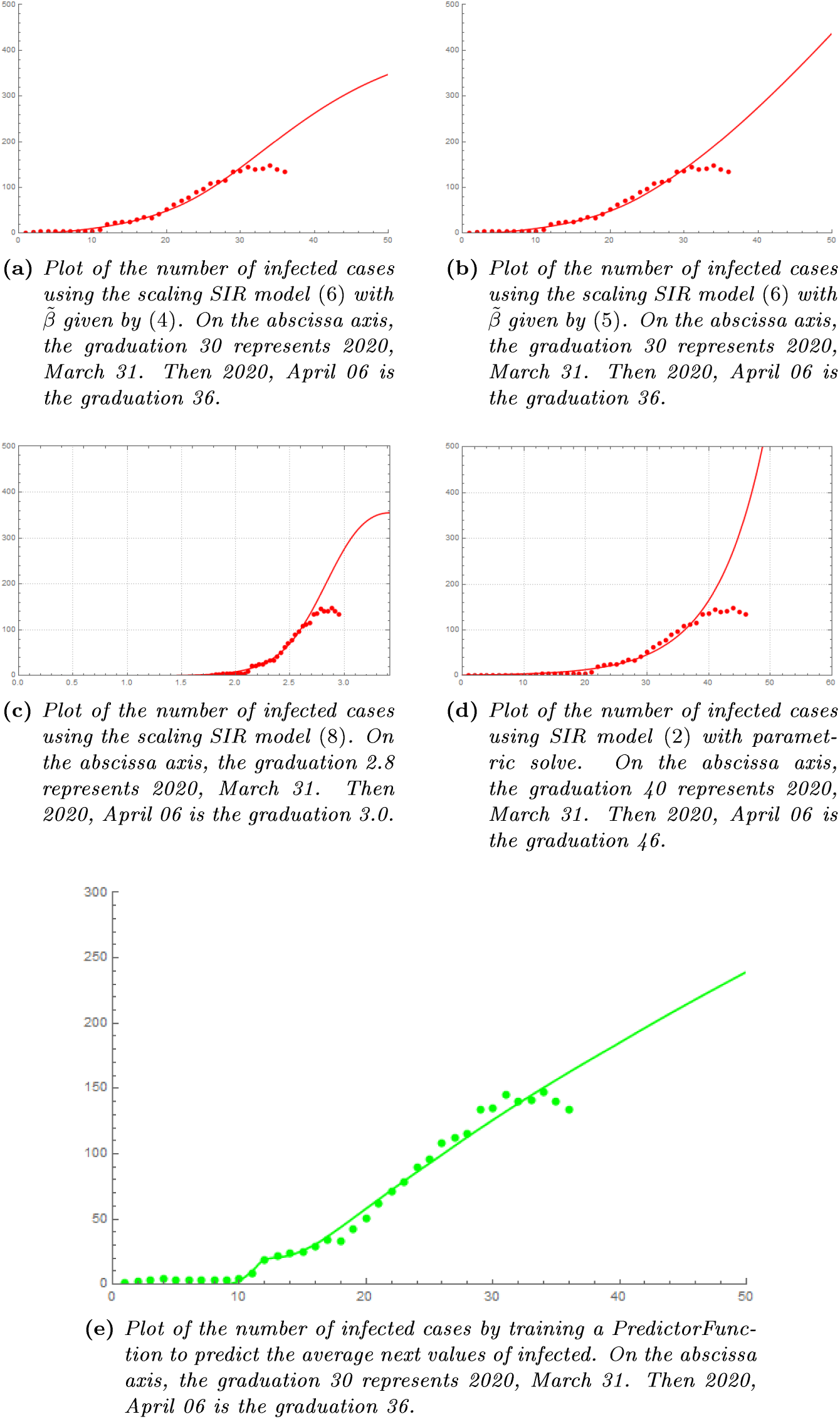
Forecasting for Senegal, using the results of the works in subsection i, ii, iii and machine learning. The data plotted (red dotted, green dotted), is given in table 1, but completed until 2020, April 06.

**Figure 11:**
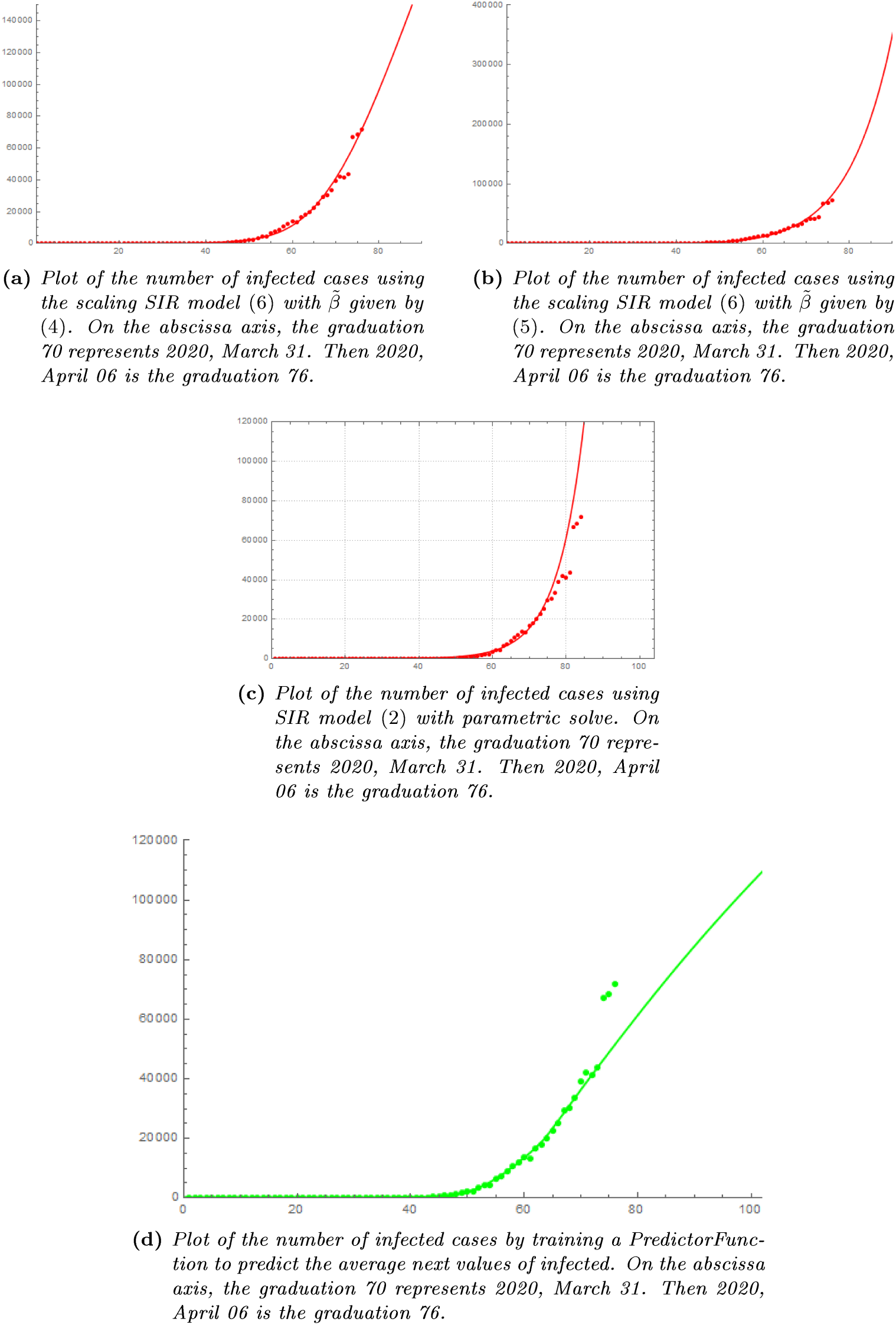
Forecasting for France, using the results of the works in subsection i,ii and iii and machine learning. The data plotted (red dotted, green dotted), is given in table 3, but completed until 2020, April 06.

## VI. Conclusion and Perspectives

In this paper, we have used the classical SIR model for fitting data and then we have done forecasting. We have also estimated the contact rate, and the average infectious period parameters of the SIR model to obtain fit. Machine learning can help in epidemiology to understand the disease, but also to study the impact or effectiveness of the anti-pandemic measures taken.

We can also study the ideal date for stopping the containment measures. Ideal in the sense that even without the measures the disease can no longer spread. We can also study the possibility of making periodic confinements to reduce the economic impact of the measures. The economy may be impacted by the evolution of the disease and measures. It would, therefore, be interesting to couple economic models with epidemiological models. Since the impact can also be long-term, scaling can be used to dimension the epidemiological-economic coupling. The impact of the environment is also to be taken into account in the models. This means studying the contamination due to the environment (air, objects, etc).

## VII. Appendix

### i. Using function fit in SIR model

From *TNI*(*t*) = *b* exp(*ct*) − *a*, we have *TNI*(*t*_0_) = *b* exp(*ct*_0_) − *a* and using (3), we obtain

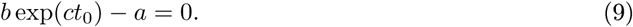

We deduce *a* = *b* exp(*ct*_0_), then 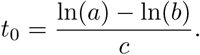

Again from (3) we have

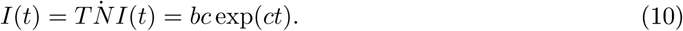

Then 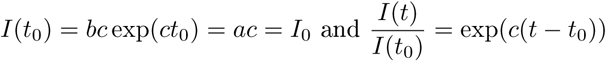. Hence, we obtain

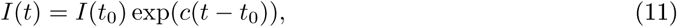

then 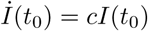.

From the second equation in the SIR model (1), we obtain at *t*_0_, 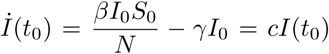. Hence, we get

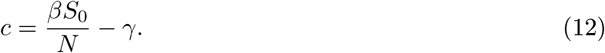

Using the third equation in the SIR model (1) yields

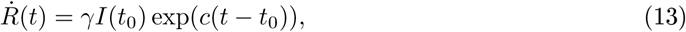

then integrating, we obtain

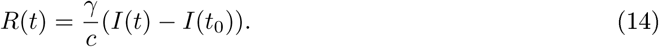

### ii. Determination of the sealed SIR

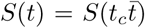, 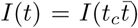 and 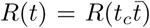, then replacing in left members of the SIR model (1), we obtain:

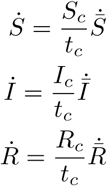

Replacing again in right members of the SIR model (1), we get:

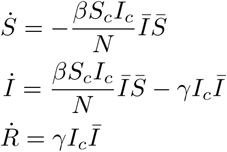

Then we have:

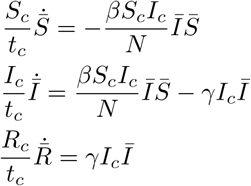

Finally, we obtain:

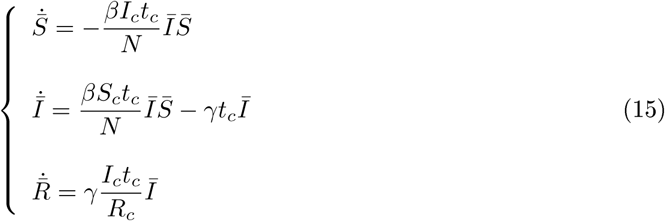

## Data Availability

I certified that all data referred to in the manuscript are available and references are given in the manuscript.

https://www.wolframcloud.com

https://github.com/antononcube/SystemModeling

http://www.sante.gouv.sn/

https://github.com/CSSEGISandData/COVID-19/issues

